# High-throughput splicing assays identify known and novel *WT1* exon 9 variants in nephrotic syndrome

**DOI:** 10.1101/2023.03.14.23287117

**Authors:** Cathy Smith, Bala Bharathi Burugula, Ian Dunn, Swaroop Aradhya, Jacob O. Kitzman, Jennifer Lai Yee

**Affiliations:** Department of Human Genetics, University of Michigan Medical School, Ann Arbor, MI 48109, USA; Department of Computational Medicine and Bioinformatics, University of Michigan Medical School, Ann Arbor, MI 48109, USA; Invitae, San Francisco, CA 94013, USA; Department of Pediatrics, Division of Nephrology, University of Michigan, Ann Arbor, MI, 48109, USA

## Abstract

Frasier Syndrome (FS) is a rare Mendelian form of nephrotic syndrome caused by variants which disrupt the proper splicing of *WT1*. This key transcription factor gene is alternatively spliced at exon 9 to produce two isoforms (“KTS+” and “KTS-”), which are normally expressed in the kidney at a ∼2:1 (KTS+:KTS-) ratio. FS results from variants that reduce this ratio by disrupting the splice donor of the KTS+ isoform. FS is extremely rare, and it is unclear whether any variants beyond the eight already known could cause FS. To prospectively identify other splicing-disruptive variants, we leveraged a massively parallel splicing assay. We tested every possible single nucleotide variant (n=519) in and around *WT1* exon 9 for effects upon exon inclusion and KTS+/- ratio. Splice disruptive variants made up 11% of the tested point variants overall, and were tightly concentrated near the canonical acceptor and the KTS+/- alternate donors. Our map successfully identified all eight known FS or focal segmental glomerulosclerosis variants and 16 additional novel variants which were comparably disruptive to these known pathogenic variants. We also identified 19 variants that, conversely, increased the KTS+/KTS- ratio, of which two are observed in unrelated individuals with 46,XX ovotesticular disorder of sex development (46,XX OTDSD). This splicing effect map can serve as functional evidence to guide the clinical interpretation of newly observed variants in and around *WT1* exon 9.

## Introduction

Variants that disrupt proper pre-mRNA splicing contribute a share of the pathogenic burden in Mendelian forms of nephrotic syndrome (NS). One NS gene sensitive to splicing disruption is *WT1*, which encodes a key genitourinary transcription factor essential for podocyte development and integrity. Its disruption results in a phenotypic spectrum including isolated NS, syndromic NS with tumors and gonadal dysgenesis, differences of sexual development, Wilms Tumor and leukemia.^1^

*WT1* undergoes alternative splicing, including in exon 9 at a pair of donors which result in protein isoforms that differ by the three amino acids, KTS. These isoforms have overlapping and district functional roles: both act as transcription factors^2^ but with partially different sequence motif specificities and gene targets.^3-6^ Normally, they are expressed in the mature kidney^7, 8^ at a ∼2:1 ratio (KTS+:KTS-). Variants which disrupt the KTS+ donor reduce this ratio and cause an extremely rare syndrome called Frasier Syndrome (FS), consisting of male gonadal dysgenesis, NS, Wilms tumor or gonadoblastoma (**Figure 1A**).^7-9^

**Figure 1.**
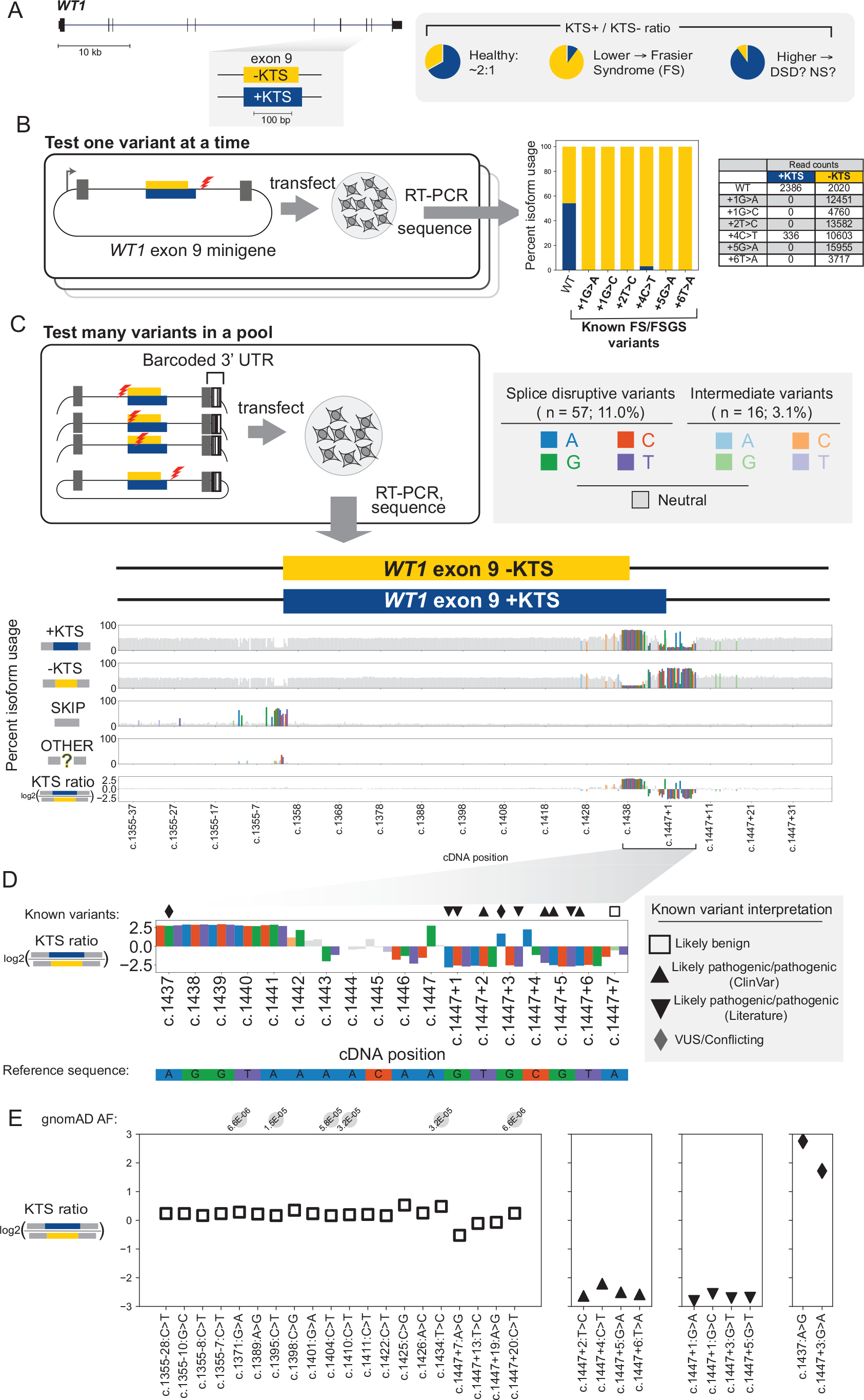
Screening for all possible splice-disruptive variants in *WT1* exon 9. **(A)** *WT1* exon 9 alternative splice forms KTS+ (blue) and KTS- (yellow). **(B)** Six known Frasier or Focal segmental glomerulosclerosis syndrome (FS/FSGS) variants tested individually by minigene assays followed by sequencing, with the percent of spliced reads from each isoform shown. **(C)** Splicing effect map for all 518 single-nucleotide variants in/around *WT1* exon 9 from a massively parallel splice assay. Each bar represents a single variant plotted by its cDNA position (x-axis), with dark shading for splice disruptive variants, light shading for intermediate ones, and gray for variants with no effect upon splicing. The first four tracks show the percent usage of KTS+, KTS-, SKIP, and OTHER isoforms. Final y-axis track shows the log2(KTS+/KTS-) ratio. **(D)** Zoom to the alternative donors showing KTS+/KTS- ratio and reference sequence. Known variants are shown above the plot with symbols denoting existing interpretation. **(E)** KTS+/KTS- ratios for variants reported in the literature or in ClinVar, grouped by interpretation, with population allele frequency shown above the plot for variants present in gnomAD.

Currently, eight variants downstream of the KTS+ splice donor are known to cause FS or focal segmental glomerulosclerosis (FSGS).^8-13^ It remains unclear if other nearby variants are similarly splice disruptive. Accurate computational prediction of variants’ splicing effects remains challenging, so we devised a massively parallel splice assay^14^ to measure the splicing effects of every possible single nucleotide variant (SNV) in or near *WT1* exon 9 and the flanking introns (N=519 variants). This identified all eight known FS/FSGS variants and nominated an additional 49 *WT1* SNVs as splice disruptive, including two patient variants with uncertain interpretations (**Table 1**). This splicing effect map can support clinical interpretation of novel *WT1* variants and improve the accuracy of genetic diagnosis.

**Table 1.**
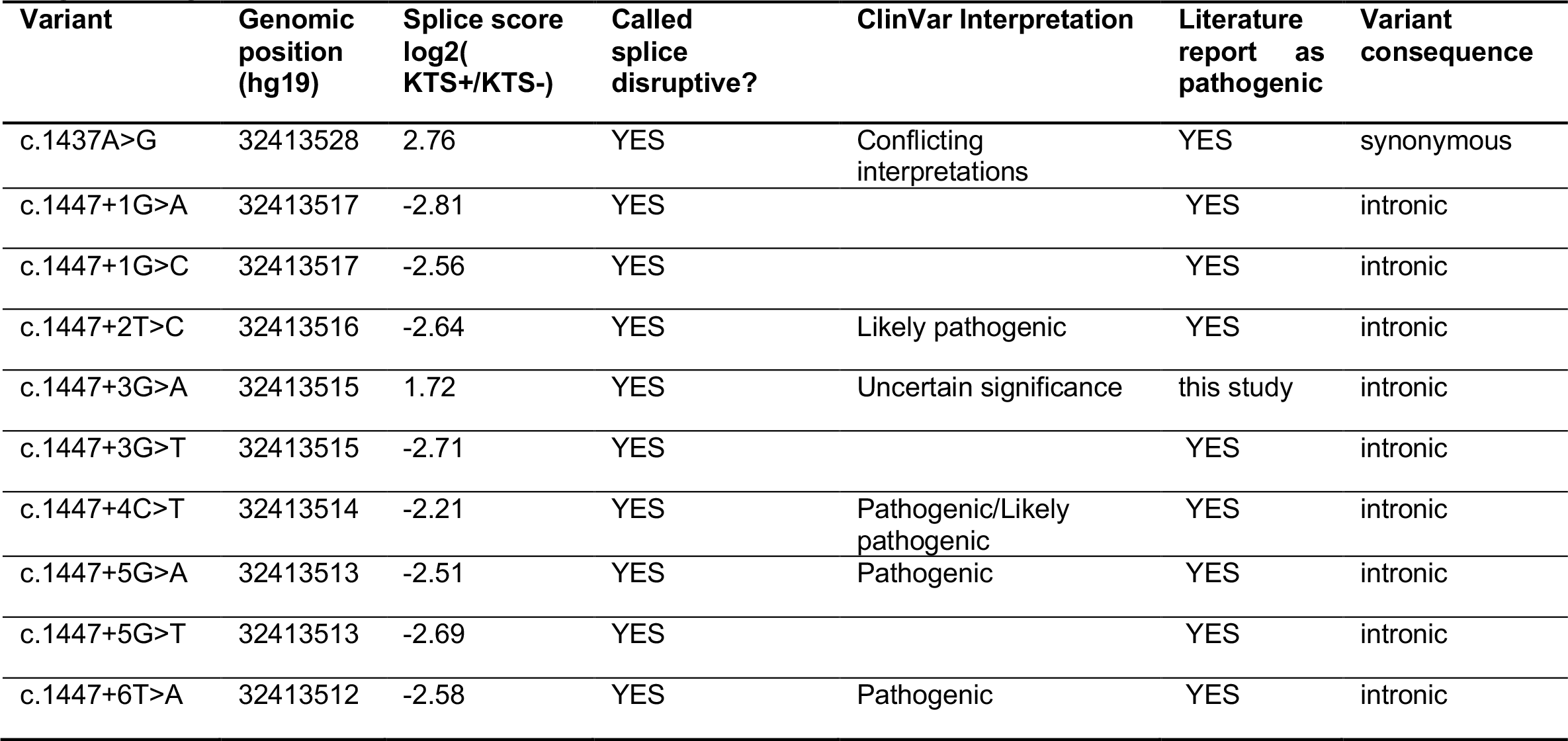
Splice assay scores for previously reported pathogenic variants for Frasier Syndrome, focal segmental glomerulosclerosis, or 46,XX OTDSD.

## Methods

### Cell culture

HEK293T and COS-7 cells were obtained from American Type Culture Collection (ATCC) and cultured in Dulbecco’s modified Eagle’s medium with high glucose, L-glutamine and sodium pyruvate (DMEM; GIBCO, Grand Island, NY, USA) containing 10% fetal bovine serum and 1% penicillin-streptomycin (10,000 U/mL) (GIBCO). Media was checked monthly for mycoplasma contamination by PCR.

### Saturation mutagenesis library construction

A *WT1* minigene construct was prepared by cloning a fragment with *WT1* exon 9 plus 414 bp of its flanking introns into the vector pSPL3 (Invitrogen, Carlsbad, CA) at the BamHI site, an intronic context flanked by a synthetic first and last exon. This construct was subjected to saturation mutagenesis as previously described^14^, targeting the full exon +/- 40 bp (173 bp). Briefly, a mutant oligonucleotide pool was designed in which each position across the targeted region was successively replaced by three other mutant bases. The pool was synthesized by Twist Biosciences (South San Francisco, CA), amplified by limited-cycle PCR, and cloned by HiFi Assembly (New England Biolabs, Ipswitch, MA) into the vector backbone linearized by inverse PCR.

### Mutant plasmid barcoding

A library of random 20mer barcode sequences were synthesized by IDT (Coralville, IA) and cloned into the downstream 3’ UTR at the MscI site by HiFi Assembly. The resulting pools of mutant *WT1* exon 9 minigenes with barcodes were transformed by electroporation into NEB 10-b E. coli, reaching library complexity of hundreds of barcoded clones per designed mutation. To enumerate the 3’UTR barcodes and identify the specific variant paired with each barcode, subassembly sequencing libraries were generated as previously described.^14, 15^

### Minigene library transfection

Mutant minigene libraries were transiently transfected into HEK293T (8 replicates) and COS-7 cells (2 replicates). At 24 hours post-transfection, cells were lysed by addition of Trizol and total RNA was purified with Direct-zol RNA Miniprep Kits (Zymo Research, Irvine, CA). A total of 3-5 ug total RNA was reverse transcribed using SuperScript III First-Strand Synthesis kit (Invitrogen) with oligo(dT)20 primer following the manufacturer’s protocol. Afterwards, spliced transcript was amplified via semi-nested PCR using outer primer pairs, first SD6 forward (5’-TCTGAGTCACCTGGACAACC-3’) and SA2 reverse (5’-ATCTCAGTGGTATTTGTGAGC-3’), and inner primer pairs, JKlab232 (5’-AGTGAACTGCACTGTGACAAGCTGC) and SA2 reverse. Indexed illumina sequencing adaptors were added by PCR and the resulting RNA-seq libraries were submitted for paired-end 150-bp sequencing on Illumina HiSeq or NovaSeq instruments.

### RNA-seq processing and splice disruption calling

RNA-seq reads were processed as previously described.^14^ Briefly, reads containing plasmid barcodes were selected with cutadapt^16^; barcodes were clustered with starcode^17^ and filtered to retain only those associated with a single-base variant. The paired, splice-informative read was aligned to the reference minigene sequence with the splice-aware read aligner STAR^18^. Custom python scripts (https://github.com/kitzmanlab/wt1_splice) were used to identify the isoform corresponding to each read: KTS+ (42.6% of all reads), KTS- (37.4%), exon 9 skipping (‘SKIP’, 19.1%), or all other isoforms (‘OTHER’; collectively, <1% of all reads). The count of reads matching each isoform was tallied per barcode, then aggregated into a per-variant, per-isoform percent by taking a read-count weighted mean of the respective percentages across the associated barcodes.

To test the significance of splice disruption, we created for each variant a null distribution by bootstrap sampling a matching number of barcodes associated with intronic variants >10 bp outside the exon boundaries. Using this null distribution, we computed z scores for the observed per-isoform usage, then used Stouffer’s method to aggregate z scores across replicates. Splice disruptive variants (SDVs) were defined as those that were (a) significant at the p<0.05 level (after Bonferroni correction for multiple testing), and either (b) had either SKIP or OTHER usage at least 20% higher than the null or (c) an isoform log-ratio (calculated as log2(KTS+/KTS-)) of >=1.5 or <= -1. Variants were defined as intermediate if they (a) passed the same significance test and had either (b) SKIP or OTHER usage at least 10% higher than the null or (c) an isoform log-ratio of >=1 or <= -.5. Results were highly correlated across replicates; all SDVs were also called disruptive at least half of the replicates when processed individually.

### Prediction of splice site strength

MaxEntScan scores^19^ for variants at the common *WT1* exon 9 acceptor, KTS-donor, and KTS+ donor were computed using the maxentpy python module (https://github.com/kepbod/maxentpy). We first computed the splice site strength for the wild-type and mutant sequences for each and took the signed difference between the variant and wild-type scores.

### Clinical variant interpretation

De-identified clinical information was obtained for carriers of WT1 exon 9 or flanking intron variants as identified in the course of clincal exome sequencing by Invitae. De-identified data were provided with institutional review board approval (WCG IRB protocol #1167406).

## Results

### Massively parallel splicing assay for *WT1* exon 9

To systematically identify splice disruptive variants, we established a minigene assay with *WT1* exon 9 and flanking introns (∼200 bp on either side). We first individually tested the wild-type sequence and six pathogenic variants near the KTS+ donor known to cause FS or FSGS (**Supplementary Table 1**). The wild-type construct showed a roughly even balance between the two isoforms (1:1.2 KTS+:KTS-ratio), while each of the known pathogenic variants abolished KTS+ usage (**Figure 1B**; **Supplementary Figure 1**). Thus, consistent with previous reports,^20^ minigenes can faithfully model *W*T1 splicing defects associated with FS and FSGS.

We next set out to test the splicing effects of every possible single nucleotide variant (SNV) in and around *WT1* exon 9 (**Figure 1C**). We applied saturation mutagenesis to create a library of all possible SNVs in the exon and for 40 bp into each flanking intron. The mutant library was tagged with random 20mers in the 3’UTR to serve as barcodes allowing for the splicing effect of each mutation to be tracked. Nearly every possible SNV was represented (518/519; 99.8%) with a high degree of internal replication (mean=79.7 barcodes per variant; **Supplemental Figure 2**).

We transfected HEK293T cells with the mutant minigene library pool and deeply sequenced the resulting spliced RNAs to quantify, for each mutation, the use of the KTS+ and KTS-isoforms, exon skipping (‘SKIP’) or all other splicing outcomes (‘OTHER’). Mutations’ effects upon isoform usage were reproducible within the HEK293T biological replicates (median pairwise Pearson’s *r=*.94) and between HEK293T replicates and a second cell line, COS-7 (median between cell line pairwise Pearson’s *r*=.93; **Supplemental Figure 3**).

The resulting map shows that, as expected, sensitivity to splicing disruption is heavily concentrated near the canonical splice sites, in particular the alternate KTS+ and KTS-donors (**Figure 1C**). Overall, of the 518 measured SNVs, only 57 (11.0%) altered splicing with an additional 16 (3.1%) having an intermediate effect on splicing (**Table 1**; **Supplemental Table 2**). Of the disruptive variants, all but one were near (+/- 15 bp) either the splice acceptor or one of the donors, consistent with disruption of those sites’ consensus motifs. The primary disruptive effect for most variants (43/57; 75.4%) was to alter the KTS+/KTS- ratio, roughly evenly split between shifting the balance towards KTS- and KTS+ (24 variants and 19 variants, respectively). A minority of variants led to complete exon skipping (n=14) or activated a cryptic acceptor 17 bp downstream of the native one (n=2), each of which would yield frameshifted transcript predicted to undergo nonsense mediated decay.

### Identification of known and novel variants disrupting KTS+ usage

We focused first on the eight known FS/FSGS variants as described in the ClinVar database or published case reports.^8-13^ All eight dramatically lowered the KTS+:KTS-balance, as quantified by log2(KTS+/KTS-) scores of -2.21 or lower **(Figure 1D, E; Table 1**). By contrast, our assay scored as neutral in all but one of the 19 variants listed in ClinVar with an interpretation of Likely Benign, as well as an additional 17 variants observed in the population database gnomAD (log2 ratio scores between -0.11 and 0.53; **Figure 1E, Supplemental Table 2**). The lone exception was c.1447+7A>G, listed in ClinVar as Likely Benign, for which we noted a very subtle shift towards KTS-(score: -0.52) for which the *in vivo* impact is unclear. Thus, pooled minigene assays effectively discriminate between known pathogenic splice disruptive variants and neutral polymorphisms.

We next asked whether this map could prospectively identify as-yet unreported variants which disrupt KTS+. We identified sixteen additional SDVs which disrupted KTS+ comparably to the known FS/FSGS variants (median log2 ratio score: -2.14; **Supplemental Table 2**), most of which were corroborated by splice site strength predictions from MaxEntScan (median MaxEntScan score=-2.40; **Supplemental Figure 4)**^19^. Among these, six were located within the codon region specific to the KTS+ isoform; four of these were synonymous variants, which as a class may be overlooked during classification. None of these is yet deposited in ClinVar nor in published reports, but based upon their disruptiveness in this assay, they represent potential novel pathogenic variants.

### Other splice disruptive outcomes

Finally, we searched this map for variants that disrupt splicing in other ways and identified nineteen variants that increased the KTS+/- ratio, opposite the effect associated with FS (**Figure 1D, E**). Notably, two have been observed in patients with 46,XX ovotesticular differences in sexual development (46,XX OTDSD). The first, c.1437A>G, is a synonymous variant recently reported^21^ as a *de novo* mutation in a patient with 46,XX OTDSD. Consistent with the strong shift towards KTS+ in our map (score: 2.76), it is predicted by MaxEntScan to weaken the KTS-donor two bases downstream **(Supplemental Figure 4)**. We observed similar effects from an additional 17 variants near the KTS-donor (median log2 ratio score: 2.83, range: 2.20-2.98). Another variant, c.1447+3G>A (log2 ratio score: 1.72), downstream of the KTS+ donor, was observed during clinical exome sequencing in a proband (age range: 10-15 y) with 46,XX OTDSD. Notably, no renal abnormalities or a history of Wlims’ tumor was reported for either of these two individuals. In contrast to the variants near the KTS-donor, this and one other variant not yet observed clinically (c.1447+4C>A) are predicted to strengthen the KTS+ donor **(Supplemental Figure 4)**, possibly leading it to outcompete its upstream counterpart. Finally, we identified a cluster of 14 variants within 26 bp of the *WT1* acceptor which led to complete skipping of exon 9, or use of an alternate cryptic acceptors, in each case leading a frameshift and premature truncation (**Supplemental Table 2**). None of those variants are yet reported in ClinVar or population databases.

## Discussion

Here, we applied a massively parallel splicing assay to systematically test the effects of every single-nucleotide variant in and near *WT1* exon 9, a hotspot of pathogenic variants for multiple genetic forms of nephrotic syndrome including Frasier Syndrome. The resulting splicing effect map correctly identified all seven known FS variants^8-11, 13^ as associated with reduced KTS+/KTS- ratio, along with another variant reported to cause isolated steroid resistant NS and FSGS.^12^ By contrast, all but one of the 36 tested variants which appear in the ClinVar database with a Likely benign interpretation, or the general healthy population database gnomAD, scored as splice-neutral, indicating this assay is highly sensitive and specific for identification of pathogenic *WT1* exon 9 splice disruption.

FS is extremely rare: in all, fewer than 200 cases have been reported, represented by only seven distinct FS variants.^8-11, 13^ Two of these seven variants (at the +4 and +5 positions) account for most of the FS case reports to date: taking the count of ClinVar submissions as a proxy for frequency, c.1447+4C>T and c.1447+5G>A together had 24 records, compared with only two records combined across the five other known FS variants. The two recurrent variants overlap the only CpG dinucleotide in the KTS+ region, and their frequency is likely explained by the ∼10-fold higher *de novo* mutation rate at germline-methylated CpGs^22^. Thus, it is reasonable to expect that there may be a tail of additional variants which are rare even within the context of this rare disorder. Indeed, our results implicate an additional sixteen SNVs as similarly decreasing the KTS+/KTS- ratio, and nominate these as new, as yet-unreported variants with the potential to cause FS/FSGS.

Our map also identified nineteen variants which increase the KTS+/KTS- ratio, either by weakening the KTS-donor or strengthening the KTS+ donor. One of these variants was previously reported^21^ in an individual with 46,XX OTDSD, and we report an additional, unrelated patient with a similar presentation carrying a different variant. Our results are consistent with these variants acting to shift the balance of WT1 isoforms toward KTS+, which is known to activate *SRY*, the master regulator of male sex determination^23^.

In some Mendelian disorders, missense or synonymous variants may alter splicing by disrupting regulatory elements beyond the canonical splice sites, termed exonic splice enhancers and silencers.^24^ Such effects have been observed by other systematic splice assays^14^ including at other *WT1* exons^25^. Here, though, we observed variants to the interior of *WT1* exon 9 had little impact upon its splicing. This suggests that *WT1* exon 9 either does not depend on exonic splice regulatory elements for its definition, or that any such elements may be robust to perturbation by single nucleotide variants.

These results may be useful in interpreting variants found in individuals who do not display every feature of FS. For instance, variants disrupting KTS+ in karyotypically female individuals (46,XX) may lead to progressive glomerulopathy, but due to the lack of gonadal dysgenesis, FS may not be suspected and *WT1* genetic testing might not be pursued.^26^ In conclusion, our systematic screen provides a lookup table of splice disruptive variants in *WT1* exon 9, circumventing the need for single variant minigene studies. The availability of functional evidence for newly observed rare variants can facilitate their resolution, lessening the burden of variant interpretation upon clinicians, and shortening the diagnostic odyssey for NS patients.

## Data Availability

All data produced in the present work are contained in the manuscript.

## Acknowledgements

We thank Ana Morales of Invitae for assistance with review of clinical records.

## Author contributions

J.O.K. and J.L.Y. devised the study. B.B., I.D., and J.L.Y. performed experiments. C.S., J.O.K., and J.L.Y analyzed the data. S.A. abstracted clinical information. C.S., J.L.Y. and J.O.K. wrote the manuscript; all authors reviewed the manuscript prior to submission.

