## Supplementary Figures 1-4 for "High-throughput splicing assays identify known and novel *WT1* exon 9 variants in nephrotic syndrome"

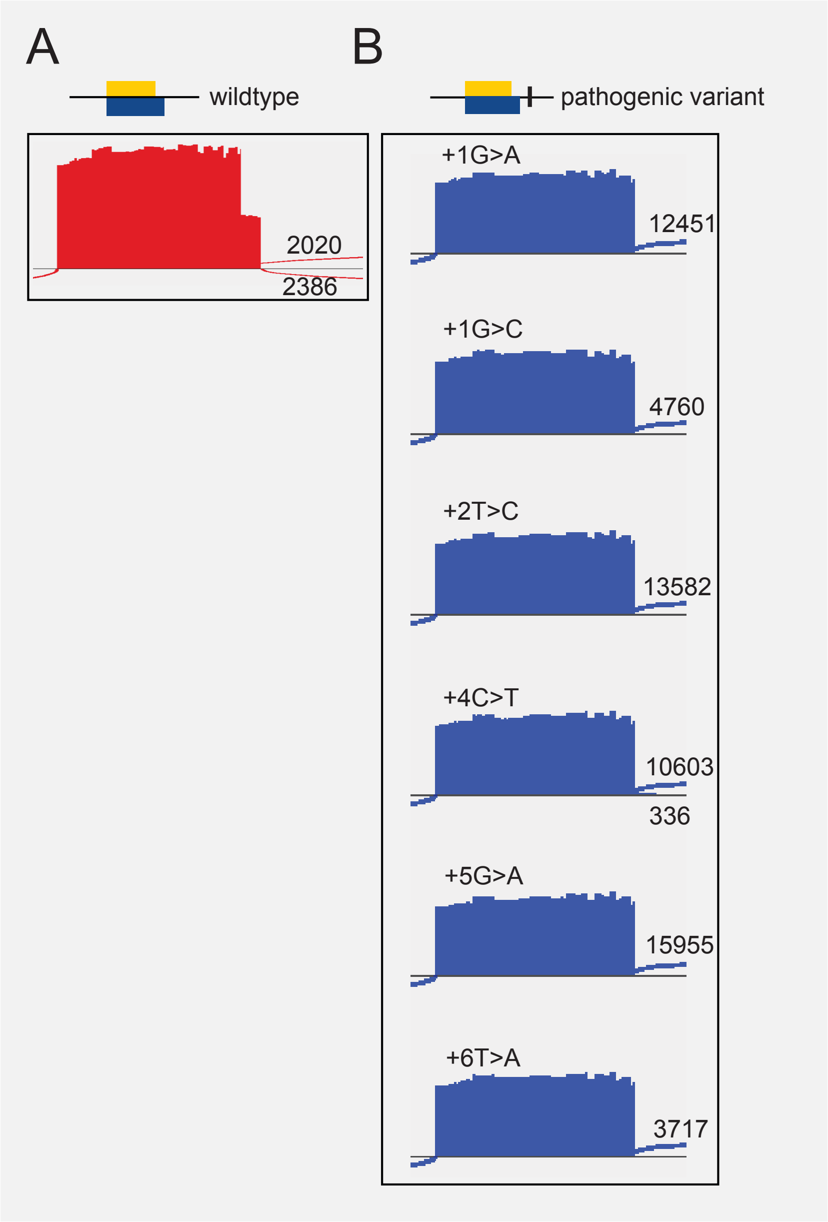


**Supplemental Figure 1** Sashimi plot from IGV showing read pileup and splice junction read counts from deep sequencing of RT-PCR products from individual minigene assays of **(A)** wildtype *WT1* exon 9 and flanking introns and **(B)** six mutant constructs each containing a different known FS/FSGS SDV. KTS- read counts are shown above each track, and KTS+ read counts (when present) are shown beneath each.

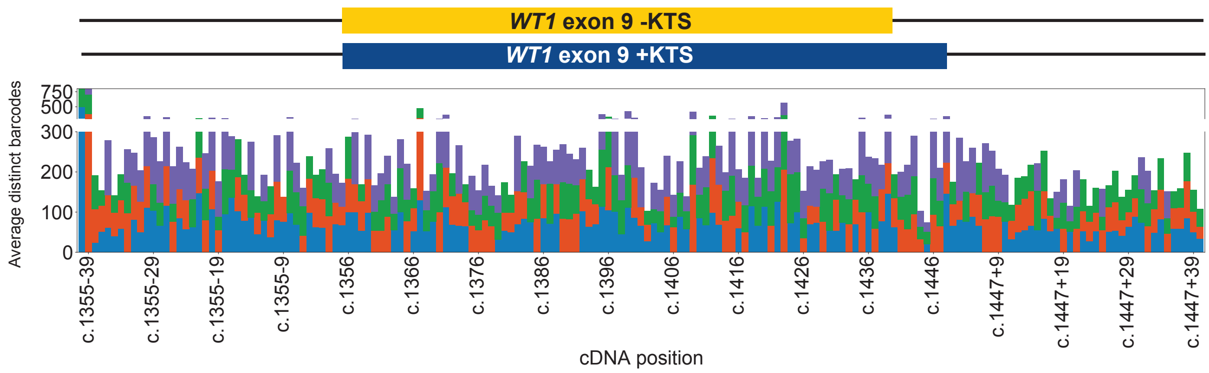


**Supplemental Figure 2.** Distinct barcode counts for each mutation (mean across replicates) are shown by position. Each shows the three different nucleotide substitutions per position.

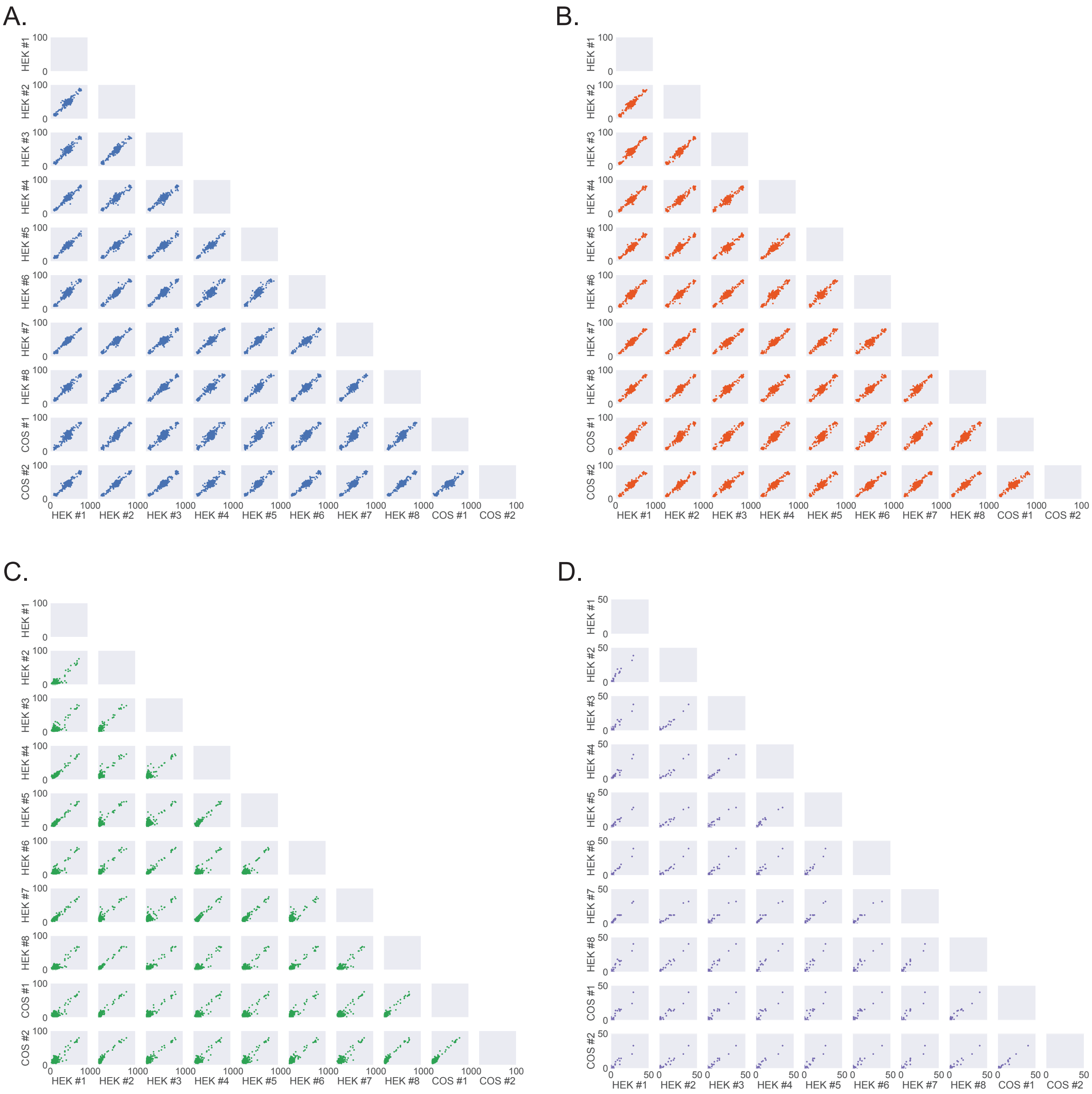


**Supplemental Figure 3.** Inter-replicate correlations of measured isoform use**. A.** KTS+ isoform usage for each replicate - different cell-types are indicated. Here, each point represents an individual variant. **B.** KTS- isoform usage for each replicate. **C.** SKIP isoform usage for each replicate. **D.** OTHER isoform usage for each replicate.

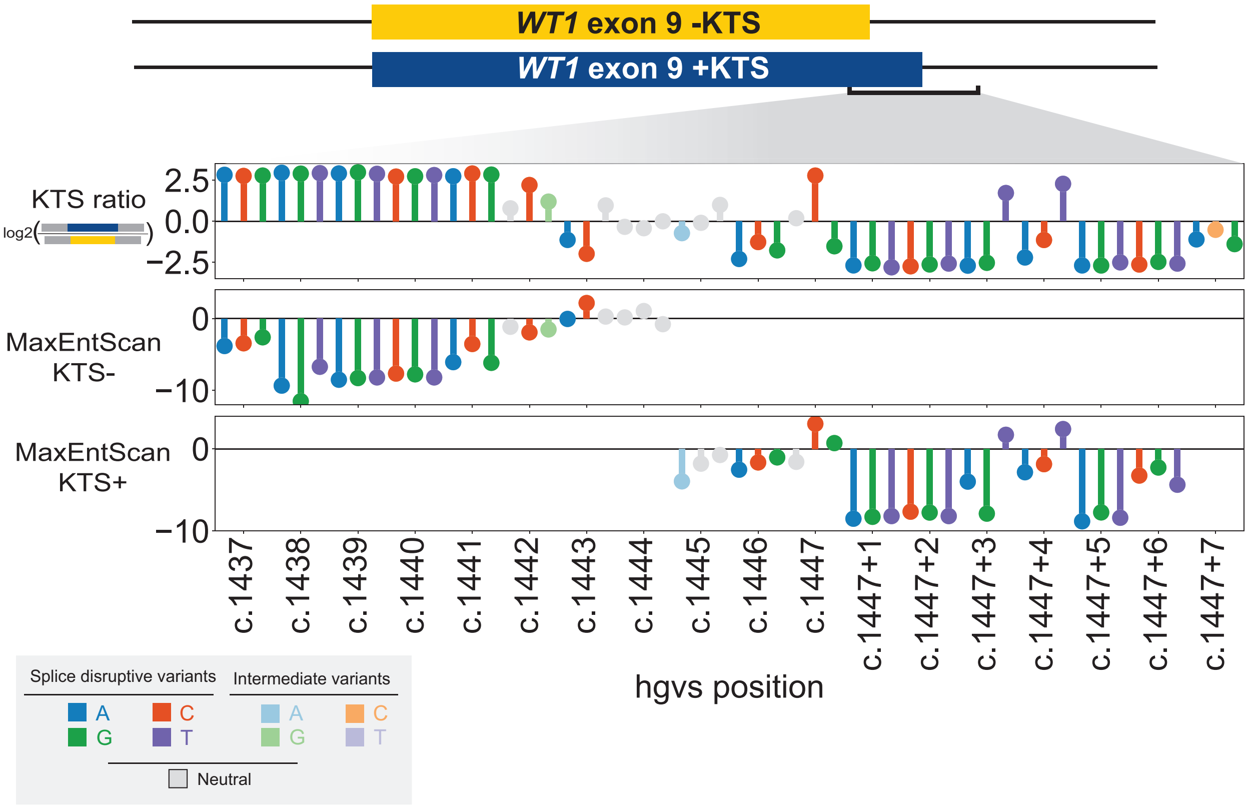


**Supplemental Figure 4.** MaxEntScan predictions of splice site strength**.** Measured splicing scores for variants surrounding the two KTS donors (top) and MaxEntScan predictions of splice site strength for the KTS- (middle) and KTS+ donors. Each lollipop represents a single variant plotted by its cDNA position (x-axis), with dark shading for variants altering the KTS ratio, light shading for intermediate ones, and gray for variants with no effect upon the KTS donors. Variants outside of the MaxEntScan scoring range have no associated lollipop.
